# *GCH1* p.Ser80Asn Confers Risk for Parkinson’s Disease in East Asian Populations

**DOI:** 10.64898/2026.06.11.26354827

**Authors:** Yi Wen Tay, Andrew Leslie Lee, Jie Ping Schee, Chin Hsien Lin, Eng King Tan, Jung Hwan Shin, Pin-Shiuan Chen, Sung-Pin Fan, Cheng-Hsuan Li, Ebonne Ng Yu Lin, Han Joon Kim, Beomseok Jeon, Sulev Koks, Kin Ying Mok, Yi Ting Lim, Mohd Salahuddin Kamaruddin, Tzi Shin Toh, Hans Xing Ding, Anis Nadhirah Khairul Anuar, Norlisah Ramli, Ignacio Juan Keller Sarmiento, Maria Teresa Periñan, Zih-Hua Fang, Lara M Lange, Kishore R. Kumar, Soraya Bardien, Joanne Trinh, Enza Maria Valente, SG10K_Health Consortium, the Global Parkinson’s Genetics Program (GP2), Peter Heutink, Katja Lohmann, Christine Klein, Niccolò E Mencacci, Shen-Yang Lim, Azlina Ahmad-Annuar, Ai Huey Tan

**Affiliations:** Department of Biomedical Science, Faculty of Medicine, University of Malaya, Kuala Lumpur, Malaysia; Division of Neurology, Department of Medicine, Faculty of Medicine, University of Malaya, Kuala Lumpur, Malaysia; Department of Neurology, National Taiwan University Hospital Taipei, Taipei, Taiwan; Duke-National University of Singapore Medical School, Singapore; Department of Neurology, National Neuroscience Institute, Singapore General Hospital, Singapore; Department of Neurology, Seoul National University Hospital, College of Medicine, Seoul National University, Seoul, Republic of Korea; Department of Neurology, National Taiwan University Hospital Bei-Hu branch, Taipei, Taiwan; Personalised Medicine Centre, Health Futures Institute, Murdoch University, Perth, Australia; Perron Institute for Neurological and Translational Science, Perth, Australia; The Hong Kong University of Science and Technology, Clear Water Bay, Kowloon, Hong Kong SAR, China; Hong Kong Center for Neurodegenerative Diseases, Hong Kong Science Park, Shatin, HKSAR, China; Department of Biomedical Imaging, Faculty of Medicine, University of Malaya, Kuala Lumpur, Malaysia; Department of Physiology, Faculty of Medicine, University of Malaya, Kuala Lumpur, Malaysia; Department of Radiology, Subang Jaya Medical Centre, Selangor, Malaysia; Ken and Ruth Davee Department of Neurology and Simpson Querrey Center for Neurogenetics, Northwestern University, Feinberg School of Medicine, Chicago, Illinois, USA; Centre for Preventive Neurology, Wolfson Institute of Population Health, Queen Mary University of London, London, United Kingdom; Unidad de Trastornos del Movimiento, Servicio de Neurología y Neurofisiología Clínica, Instituto de Biomedicina de Sevilla, Hospital Universitario Virgen del Rocío/Consejo Superior de Investigaciones Científicas (CSIC)/Universidad de Sevilla, Seville, Spain; DataTecnica LLC, Washington, DC 20037, USA; Laboratory of Neurogenetics, National Institute on Aging, National Institutes of Health, Bethesda, MD, USA; Institute of Neurogenetics, University of Luebeck, Luebeck, Germany; Translational Neurogenomics Group, ANZAC Research Institute, Sydney Local Health District and Faculty of Medicine and Health, The University of Sydney, Sydney, New South Wales, Australia; Garvan Institute of Medical Research, Sydney, NSW 2010, Australia; School of Clinical Medicine, UNSW Medicine & Health, University of New South Wales, Kensington, NSW, Australia; Division of Molecular Biology and Human Genetics, Department of Biomedical Sciences, Faculty of Medicine and Health Sciences, Stellenbosch University, Cape Town, South Africa; South African Medical Research Council/Stellenbosch University Genomics of Brain Disorders Research Unit, Cape Town, South Africa; Neurogenetics Research Center, IRCCS Mondino Foundation, 27100 Pavia, Italy; Department of Molecular Medicine, University of Pavia, 27100 Pavia, Italy

**Keywords:** *GCH1*, East Asian, Parkinson’s disease

## Abstract

**Introduction:** *GCH1* has been implicated in Parkinson’s disease (PD), but its risks variants and associations are not well defined.

**Objectives:** To investigate the clinical relevance and PD risk associated with the *GCH1* p.Ser80Asn variant.

**Methods:** We first identified a segregating *GCH1* p.Ser80Asn variant in a Malaysian Chinese PD family via whole genome sequencing (WGS). We assessed its risk association using multi-ancestry WGS data from the Global Parkinson’s Genetics Program (GP2) (n=22,372_PD_ vs n=8,826_Controls_) and meta-analysis of East Asian (EAS) cohorts (n=4,712_PD_ vs 38,733_Controls_). Clinico-demographic details of affected variant carriers were collated.

**Results:** The *GCH1* p.Ser80Asn variant was enriched in GP2 EAS PD populations (n=9/2,757; 0.33%) but not detected in other ancestries. Meta-analysis revealed increased PD risk in EAS populations (odds ratio:5.1; 95%CI:2.3–10.7; p=2.89×10^−5^). Affected carriers (mean age at onset:56.3±12.5 years) had additional occurrence of dystonia, while dementia was rare.

**Conclusions:** The *GCH1* p.Ser80Asn variant is a rare, EAS-enriched risk variant for PD.

## Introduction

Guanosine triphosphate cyclohydrolase 1 (GTPCH-1), encoded by the *GCH1* gene, is a rate-limiting enzyme in tetrahydrobiopterin synthesis, a key cofactor for dopamine production. Heterozygous *GCH1* variants are the most frequent cause of dopa-responsive dystonia (DRD).^1^ Meanwhile, the contribution of *GCH1* to PD has long been debated. Several common non-coding *GCH1* variants have been identified as PD risk loci in large genome-wide association studies (GWAS),^2-4^ including the rs11158026 variant,^5-8^ but recent studies reported inconsistent findings with potential geographical/population differences in *GCH1* risk associations.^9-16^ Notably, a significant burden of deleterious *GCH1* variants was identified in early-onset or familial PD.^14^ Segregation of rare *GCH1* variants with both DRD and PD within families, together with the identification of likely pathogenic *GCH1* variants in PD patients, further support its role as a PD-relevant gene.^13,17-19^

Here, we describe a Malaysian Chinese family carrying the *GCH1* p.Ser80Asn variant, and investigate its association with PD risk across large multi-ancestral datasets from the Global Parkinson’s Genetics Program (GP2) and East Asian cohorts. We further characterize its phenotypic spectrum through a systematic review of published variant carriers.

## Methods

This study was approved by the Universiti Malaya Medical Centre (UMMC) Medical Research Ethics Committees (No. 20191010-7917). All participants provided written informed consent. Detailed methods are provided in Supplementary Methods.

### Clinical and genetic screening of proband and family

The proband with his family (n=4; Figure 1A) was recruited from the Neurology Clinic, UMMC, Kuala Lumpur, Malaysia. Detailed assessment of all subjects was performed by a movement disorders neurologist (AHT). The proband underwent whole genome sequencing (WGS)^20^ and multiplex ligation-dependent probe amplification analysis. Candidate variant was validated using Sanger sequencing.

**Figure 1:**
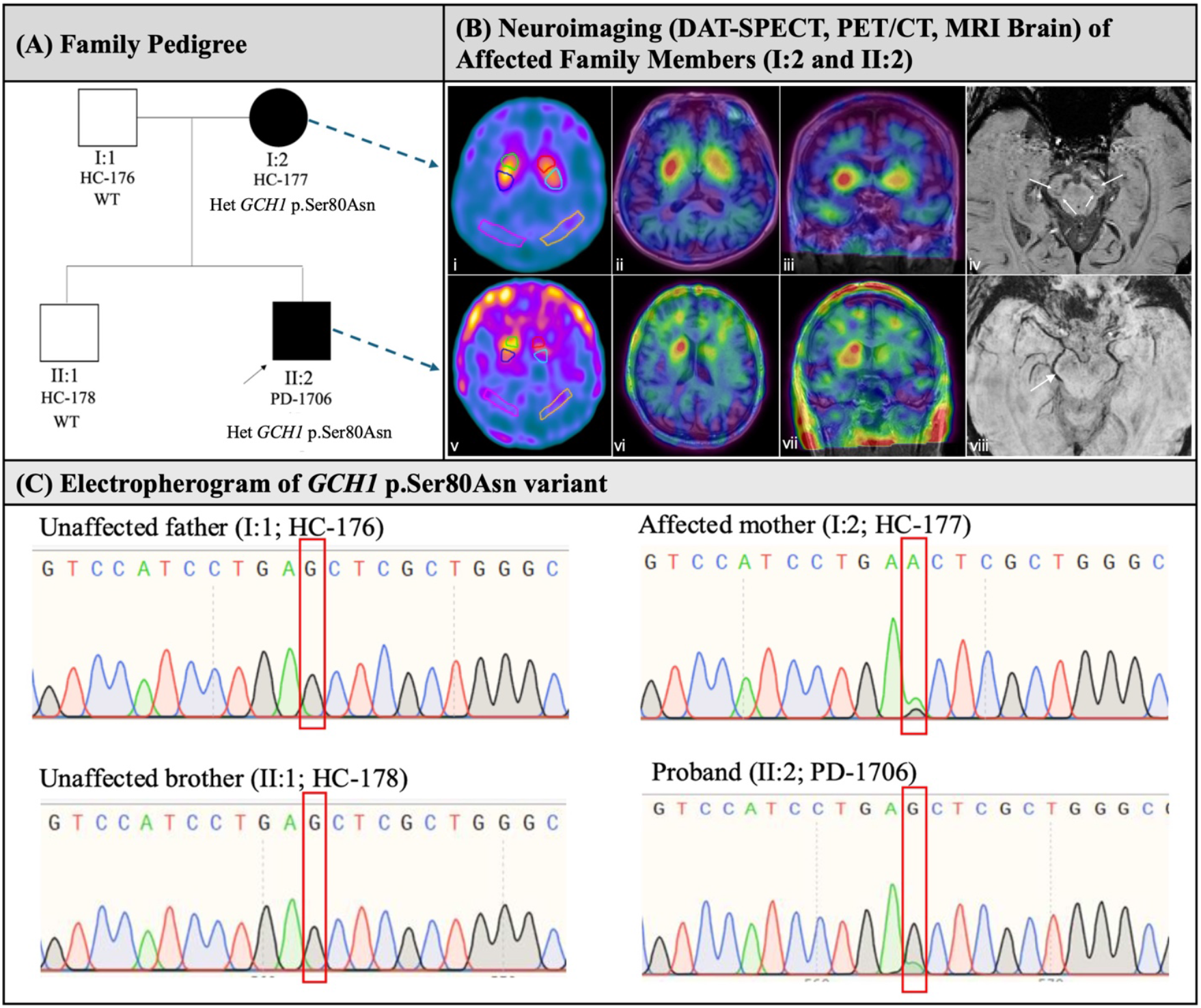
Pedigree of a Malaysian Chinese family carrying a segregating *GCH1* p.Ser80Asn variant and the clinico-radiological findings. (A) The proband (II:2) and his affected mother (I:2) were found to carry the single heterozygous *GCH1* p.Ser80Asn variant, whereas his unaffected father and brother were negative for the variant. (B) Nigrostriatal neurodegeneration is observed in both the mother (I:2) and the proband (II:2), evidenced by reduced tracer uptake in DaT-SPECT (i, v) and 99mTc-TRODAT-1 SPECT/MRI fusion images (ii, iii, vi, vii). The mother exhibits mild, primarily unilateral loss (i-iii). In contrast, the proband shows marked bilateral involvement of the putamen (v-vii), worse on the left. SWI sequence images in the axial plane (iv, viii) show preserved bilateral “swallow tail” sign in the mother (iv, white arrows), while there is a loss of the left swallow tail sign and nigrosome 1 hyperintensity, but preserved right swallow tail sign in the proband (viii, white arrow). DaT=Dopamine Transporter; DaT-SPECT=Dopamine Transporter Single-Photon Emission Computed Tomography; MRI=Magnetic Resonance Imaging; PET/CT=Positron Emission

### Risk association and haplotype analyses

Following the discovery of the segregating *GCH1* p.Ser80Asn variant in this family, we analyzed WGS data from GP2 Release 11 (DOI:10.5281/zenodo.17753486) (n=24,508) to determine its carrier frequency across 11 ancestries (Supplementary Table 1). Variant carriers were also screened for pathogenic/likely pathogenic variants in other PD-related genes (Supplementary Table 2).

Additionally, we analysed the carrier frequency in two other Asian populations (i.e., the EAS whole exome sequencing study (EAS-WES)^21^ and the Singaporean whole genome-based SG10K_Health^22^ project) (n=43,445). Allele frequencies between cases and controls were compared using two-tailed Fisher’s exact test, with calculation of odds ratio (OR) and confidence intervals (CI).

To assess a shared haplotype among carriers, chromosome 14 WGS data were phased using Beagle v5.4^23^ and analysed for identity-by-descent segments (≥2cM) using hap-ibd,^24^ complemented by analyses of flanking microsatellite and SNP markers.

### Literature search of *GCH1* p.Ser80Asn cases

A PubMed search was conducted using the terms (“GCH1” OR “GTP cyclohydrolase 1”) AND (dystonia OR “dopa-responsive dystonia” OR “DRD” OR “parkinson” OR “Parkinson’s disease”), for publications until 31st December 2025. Data on all published *GCH1* p.Ser80Asn carriers were extracted.

## Results

### Clinical and genetic findings of the proband and his family

The proband, a Malaysian Chinese male (II:2, Figure 1A), was diagnosed with early-onset PD (EOPD) ranging 40-50 years. He developed motor response complications at seven years after onset, and required high levodopa equivalent daily dose (LEDD) of 950mg/day. His mother (I:2, Figure 1A), was incidentally found, as part of research evaluation, to have mild parkinsonian features at age ranging 70-80 years and remained stable on low-dose levodopa (LEDD: 100mg/day). 99mTc-TRODAT-1 SPECT imaging (Figure 1B) confirmed nigrostriatal dopaminergic denervation in both individuals, with the proband exhibiting marked bilateral putaminal involvement compared with his mother’s mild, primarily unilateral loss. Detailed clinical progression, including rating-scales and videos, are provided in Supplementary Materials. Both the father (I:1, Figure 1A) and elder brother (II:1, Figure 1A) were neurologically normal.

The proband was found to carry a heterozygous *GCH1* p.Ser80Asn (c.239G>A; NM_000161.3) variant. This variant has a CADD score of 21.7, and fulfilled the American College of Medical Genetics and Genomics (ACMG) criteria of PM1, PP2, PM2, and BP6. It was classified as a variant of unknown significance by Franklin.^25^ Screening for other pathogenic/likely pathogenic variants in other PD-related genes was negative.

His affected mother also carried the same variant, whereas his unaffected father and brother did not (Figure 1C), suggesting a segregation with disease and an autosomal dominant mode of inheritance.

### Carrier frequency and risk ascertainment in multi-ancestry cohorts

In the GP2 R11 WGS dataset (n=22,372_PD_; n=3,578_other neurological disorders_; n=8,826_Controls_), the *GCH1* p.Ser80Asn variant was found exclusively in EAS-ancestry individuals (n=9/2,757 PD patients and n=1/1,020 control; minor allele frequency [MAF]=0.16% in cases and 0.05% in controls). Among the unrelated carriers, 5 were from Malaysia (n=5/1,516, 0.33%), 3 from Taiwan (n=3/575, 0.52%), and 1 from South Korea (n=1/298, 0.34%), as well as one unrelated EAS control from Malaysia (Supplementary Table 1). None of the carriers had pathogenic/likely pathogenic variants (including CNVs for *PRKN* and *SNCA*) in known PD genes,^26^ except for one Malaysian patient (Subject 6) who had a pathogenic *GBA1* variant (c.762-1G>C).

In independent cohorts, 4/1,955 Singaporean Chinese PD patients carried the variant (0.20%), whereas no carriers were identified among 5,512 EAS-WES controls or 9,770 EAS controls from the SG10K_Health dataset. In gnomAD (v4.1.1), the *GCH1* p.Ser80Asn variant had a global allele frequency of 0.001%, and was observed only in individuals of EAS ancestry (n=20/22,431; MAF=0.04%), with no homozygotes identified.

Meta-analysis of the EAS PD patients from GP2 and the Singapore cohort (n=4,712, 13 carriers; MAF=0.14%) versus EAS controls from GP2, EAS-WES, SG10K_Health, and gnomAD datasets (n=38,733; 21 carriers; MAF=0.03%) revealed an OR of 5.1 (95%CI=2.3-10.7, p=2.89×10^−5^). Taken together, our findings suggest that *GCH1* p.Ser80Asn variant is a rare but significant PD risk variant in EAS populations, providing evidence consistent with ACMG PS4 criterion (enrichment in affected individuals compared with controls), thereby supporting its reclassification of variant classification to “likely pathogenic”.

### Haplotype analyses

Haplotype analyses revealed no shared disease-associated haplotype among the *GCH1* p.Ser80Asn carriers in EAS cohort (Supplementary Table 3).

### Clinico-demographic characteristics of affected carriers

Of the 323 PubMed records retrieved, three publications reported a total of five *GCH1* p.Ser80Asn variant carriers (one patient with DRD, three with PD, and one unaffected family member), all of Chinese ancestry.^27-29^

The clinico-demographic features of newly identified and published variant carriers are summarized in Table 1. The mean age at PD onset was 56.3±12.5 years (range:40-77); 38.5% (n=5/13) had EOPD.^30^ Only two probands reported a positive family history of PD; none reported a family history of dystonia. Phenotypically, we noted a high frequency of resting tremor (n=10/13, 76.9%), and a notable presence of focal or multifocal dystonia in six patients (46.2%). The distribution of dystonia was heterogeneous, from isolated foot dystonia, blepharospasm or cervical dystonia, to multi-focal involvement of the trunk and limbs.

**Table 1:**
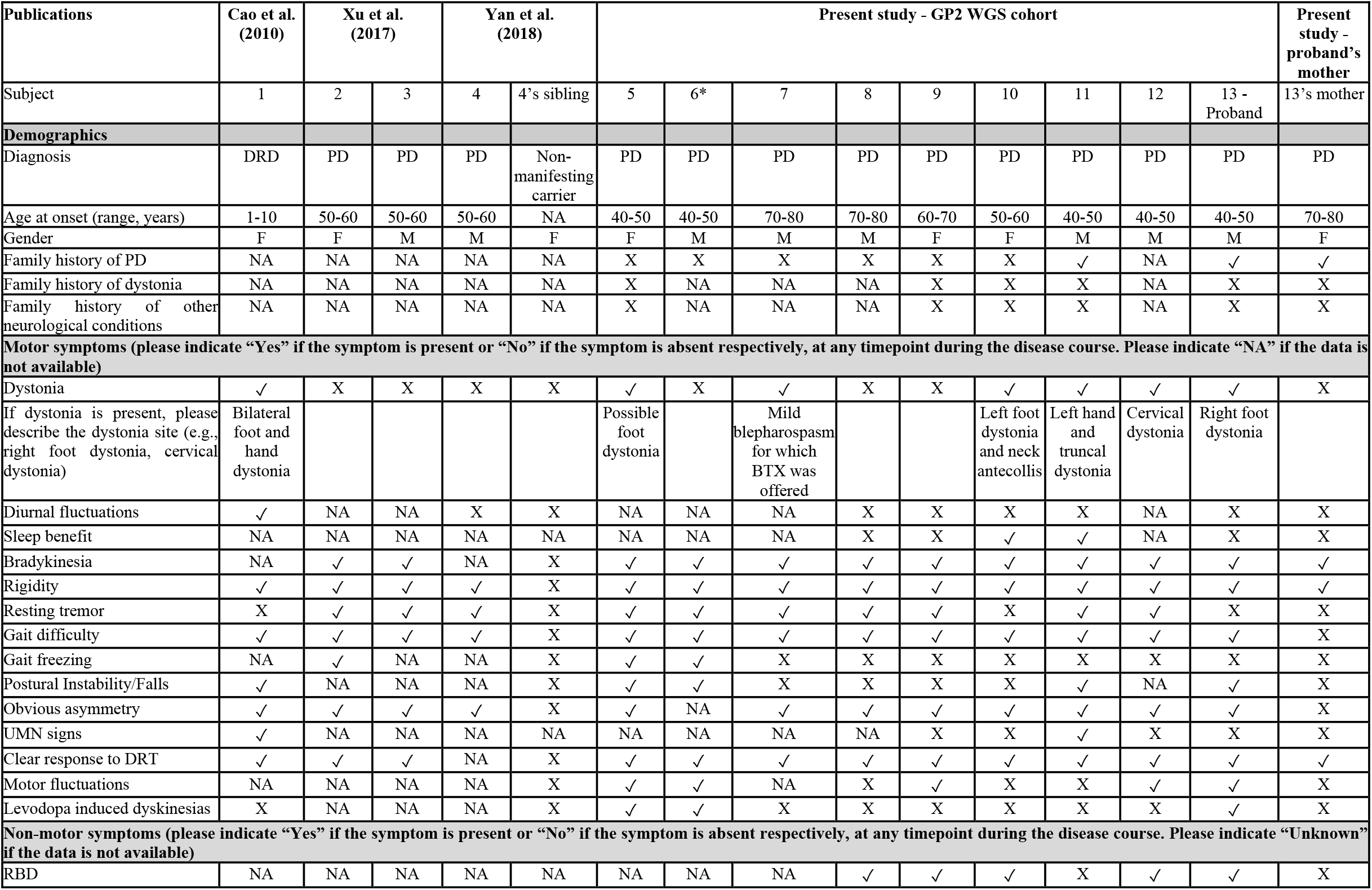
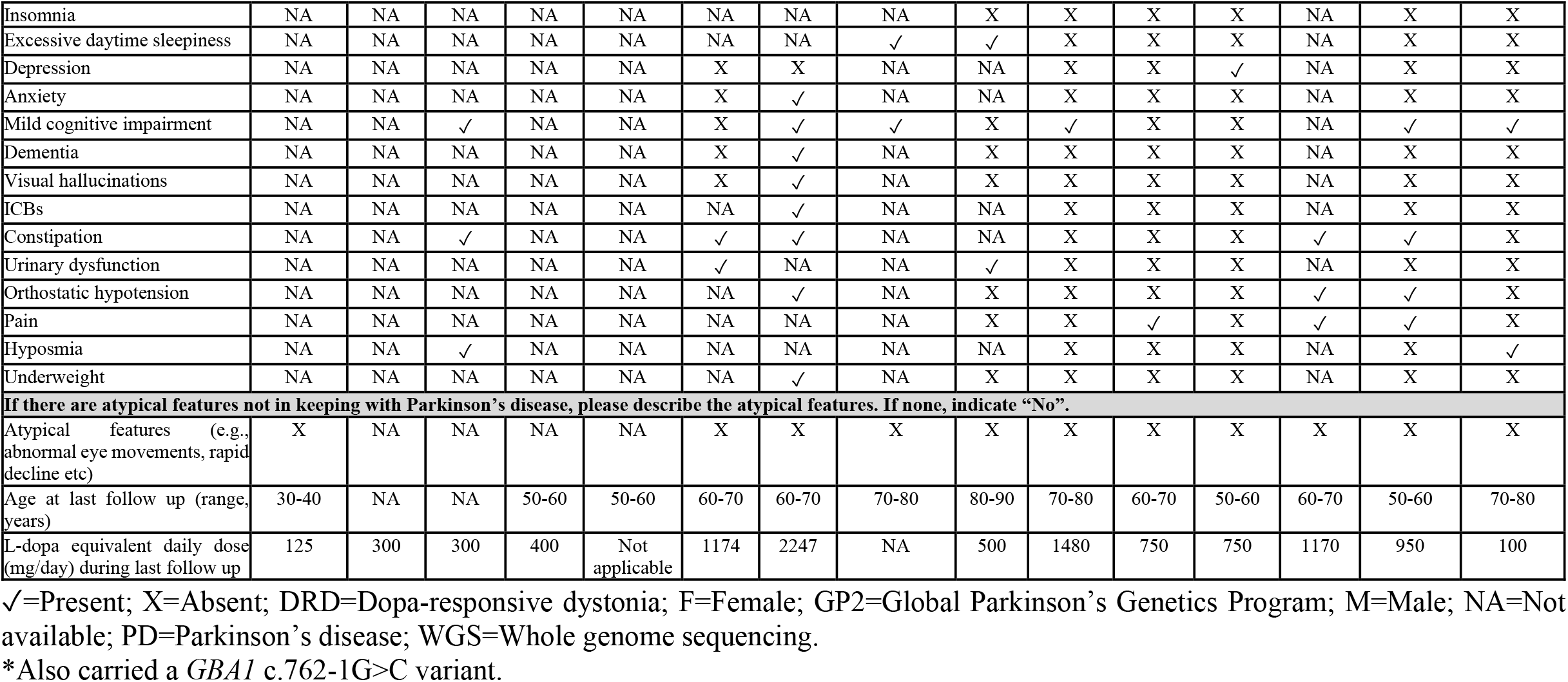
Clinico-demographic characteristics of East Asian carriers of the *GCH1* p.Ser80Asn variant from published reports and the present study.

Most patients had a favorable response to dopaminergic therapy (LEDD: 300-1480mg/day). Motor fluctuations were present in 55.6% (n=5/9), with levodopa-induced dyskinesia in 30% (n=3/10). Non-motor symptoms were common, including RBD (71.4%, n=5/7), mild cognitive impairment (60%, n=6/10), constipation (55.6%, n=5/9), hyposmia (33.3%, n=2/6), and urinary dysfunction (28.6%, n=2/7). Dementia was reported 13 years after disease onset in Subject 6 who co-carried the *GBA1* c.762-1G>C variant. None reported significant diurnal variation in symptoms, although two patients reported sleep benefit.

## Discussion

Our study identified *GCH1* p.Ser80Asn as a rare risk variant for PD in EAS populations. We demonstrated co-segregation of this variant with levodopa-responsive parkinsonism in a small Malaysian Chinese pedigree, accompanied by neuroimaging evidence of dopaminergic denervation, consistent with PD. Meanwhile, large-scale multi-ancestry screening across >50,000 individuals revealed that the variant was exclusive to EAS populations with significant enrichment in PD cases (OR 5.3), allowing reclassification of the variant as likely pathogenic. Comprehensive review of variant carriers highlighted a low prevalence of dementia and notable presence of focal or multifocal dystonia among affected carriers, as well as incomplete penetrance and variable expressivity within families.

To date, >150 *GCH1* variants have been reported.^1,31^ In our study, the presence of the p.Ser80Asn variant in an asymptomatic sibling (Table 1) and in one control, as well as the absence of a positive family history in most carriers, suggests incomplete penetrance, in keeping with observations reported for other *GCH1* variants.^13,32,33^ Although a higher penetrance has been reported in females than males with *GCH1*-related DRD,^34,35^ this pattern was not seen in our PD cohort. We also observed variable expressivity among carriers of p.Ser80Asn. For example, while our Malaysian Chinese proband had EOPD and a more aggressive disease course with motor response complications, his mother carrying the same variant displayed a late onset and indolent progression. Additionally, the *GCH1* p.Ser80Asn variant has been reported in one patient with DRD (age of onset ranging 1-10 years).^27^ Beyond DRD and PD, rare *GCH1* variants have also been reported in patients with hereditary spastic paraplegia, underscoring the complex genotype-phenotype relationships of this gene.^36^

The p.Ser80Asn variant lies near the start of the cyclohydrol domain, which mediates the catalytic activity of GCH1.^37^ Variants in this domain reduce GCH1 activity and impair tetrahydrobiopterin-dependent dopamine synthesis. >80–90% of rare missense *GCH1* variants reported in dystonia and PD localize here, suggesting that phenotypic variability among *GCH1* carriers is unlikely to be solely explained by variant location or enzyme level.^28^ The mechanism whereby *GCH1* variants predispose to PD remains unclear with lack of experimental studies, but several hypotheses are plausible. Firstly, chronic dopamine deficiency and/or associated compensatory responses since childhood may increase nigral vulnerability to ageing, environmental, or genetic stressors, leading to neurodegeneration.^13^ Alternatively, this neurochemical deficiency may also lower the threshold at which nigral cell loss results in parkinsonian symptoms, analogous to the pathomechanism of drug-induced parkinsonism.^38^ Finally, the role of other cellular or molecular (e.g., non-dopaminergic) pathways cannot be excluded.

This study has several limitations. Firstly, our pedigree was small, precluding definitive conclusions about co-segregation with disease and penetrance. Larger multi-center cohorts with deep phenotyping and longitudinal follow-up will be essential to refine estimates of age-dependent penetrance and expressivity, and the contribution of environmental or genetic modifiers.^39^ Secondly, functional experiments^40^ (e.g., measurements of GTPCH-1 activity and tetrahydrobiopterin level, interactions with dopaminergic stress or α-synuclein pathology) were not performed. Future investigations into the downstream effects of this variant are warranted and may reveal new mechanistic insights for disease-modifying strategies in *GCH1*-associated PD.

In conclusion, our study identified *GCH1* p.Ser80Asn as a rare, East Asian risk variant for PD. This discovery highlights the importance of family studies and ancestral diversity in genetic discovery and suggests that rare, moderate-effect variants may contribute to PD risk architecture with translational potential in underrepresented populations.

## Acknowledgement and author roles

The authors gratefully acknowledge the family for their consent and participation in this study, including the publication of the videos online. This project was supported by the Global Parkinson’s Genetics Program (GP2; https://gp2.org). GP2 is funded by the Aligning Science Across Parkinson’s (ASAP) (https://ror.org/03zj4c476) initiative and implemented by The Michael J. Fox Foundation for Parkinson’s Research (MJFF) (https://ror.org/03arq3225). For a complete list of GP2 members see https://doi.org/10.5281/zenodo.7904831. This research was also supported in part by the Intramural Research Program of the National Institutes of Health (NIH). The contributions of the NIH author(s) are considered Works of the United States Government. The findings and conclusions presented in this paper are those of the author(s) and do not necessarily reflect the views of the NIH or the U.S. Department of Health and Human Services. In addition, this study made use of data generated as part of the Singapore National Precision Medicine program funded by the Industry Alignment Fund (Pre-Positioning) (IAF-PP: H17/01/a0/007). This study made use of data / samples collected in the following cohorts in Singapore: (1) The Health for Life in Singapore (HELIOS) study at the Lee Kong Chian School of Medicine, Nanyang Technological University, Singapore (supported by grants from a Strategic Initiative at Lee Kong Chian School of Medicine, the Singapore Ministry of Health (MOH) under its Singapore Translational Research Investigator Award (NMRC/STaR/0028/2017) and the IAF-PP:H18/01/a0/016); (2) The Growing up in Singapore Towards Healthy Outcomes (GUSTO) study, which is jointly hosted by the National University Hospital (NUH), KK Women’s and Children’s Hospital (KKH), the National University of Singapore (NUS) and the Singapore Institute for Clinical Sciences (SICS), Agency for Science Technology and Research (A*STAR) (supported by the Singapore National Research Foundation under its Translational and Clinical Research (TCR) Flagship Programme and administered by the Singapore Ministry of Health’s National Medical Research Council (NMRC), Singapore - NMRC/TCR/004-NUS/2008; NMRC/TCR/012-NUHS/2014. Additional funding is provided by SICS and IAF-PP H17/01/a0/005); (3) The Singapore Epidemiology of Eye Diseases (SEED) cohort at Singapore Eye Research Institute (SERI) (supported by NMRC/CIRG/1417/2015; NMRC/CIRG/1488/2018; NMRC/OFLCG/004/2018); (4) The Multi-Ethnic Cohort (MEC) cohort (supported by NMRC grant 0838/2004; BMRC grant 03/1/27/18/216; 05/1/21/19/425; 11/1/21/19/678, Ministry of Health, Singapore, National University of Singapore and National University Health System, Singapore); (5) The SingHealth Duke-NUS Institute of Precision Medicine (PRISM) cohort (supported by NMRC/CG/M006/2017_NHCS; NMRC/STaR/0011/2012, NMRC/STaR/ 0026/2015, Lee Foundation and Tanoto Foundation); and (6) The TTSH Personalised Medicine Normal Controls (TTSH) cohort funded (supported by NMRC/CG12AUG17 and CGAug16M012). The views expressed are those of the author(s) are not necessarily those of the National Precision Medicine investigators, or institutional partners. We thank all investigators, staff members and study participants who made the National Precision Medicine Project possible.

## Author Roles

(1) Research Project: A. Conception and Design, B. Data Acquisition, C. Data Analysis; (2) Statistical Analysis: A. Design, B. Execution, C. Review and Critique; (3) Manuscript Preparation: A. Drafting the Manuscript and/or Figures, B. Review and Critique; C. Final Approval.

YWT: 1A, 1B, 1C, 2A, 2B, 2C, 3A, 3B, 3C

ALL: 1B, 1C, 3A, 3C

JPS: 1B, 3A, 3C

CHL: 1B, 2C, 3B, 3C

EKT: 1B, 2C, 3B, 3C

JHS: 1B, 2C, 3B, 3C

P-SC: 1B, 3B, 3C

S-PF: 1B, 3B, 3C

C-HL: 1B, 3B, 3C

ENYL: 1B, 3B, 3C

HJK: 1B, 3B, 3C

BJ: 1B, 3B, 3C

SK: 1B, 3B, 3C

KYM: 1B, 3B, 3C

YTL: 1B, 3B, 3C

MSK: 1B, 3B, 3C

TST: 1B, 3B, 3C

HXD: 1B, 3B, 3C

ANKA: 1B, 3B, 3C

NR: 1B, 3B, 3C

IJKS: 2C, 3B, 3C

MTP: 2C, 3B, 3C

Z-HF: 1B, 3B, 3C

LML: 1B, 2C, 3B, 3C

KRK: 2C, 3B, 3C

SB: 2C, 3B, 3C

JT: 2C, 3B, 3C

EMV: 2C, 3B, 3C

PH: 2C, 3B, 3C

KL: 2C, 3B, 3C

CK: 2C, 3B, 3C

NEM: 1A, 2C, 3B, 3C

SYL: 1B, 1C, 2C, 3B, 3C

AAA: 1B, 1C, 2C, 3B, 3C

AHT: 1A, 1B, 1C, 2A, 2C, 3A, 3B, 3C

## Data Availability

Data used in the preparation of this article were obtained from the Global Parkinson’s Genetics Program (GP2; https://gp2.org). Specifically, we used Tier 2 data from GP2 release 11 (10.5281/zenodo.17753486). GP2 data can be accessed through AMP PD (https://amp-pd.org). All code generated for this article, and the identifiers for all software programs and packages used, are available on GitHub (https://github.com/GP2code/EAS_GCH1_pS80N) and were given a persistent identifier via Zenodo (10.5281/zenodo.20562807).

### Ethical Compliance Statement

Institutional review board approval was obtained from the Universiti Malaya Medical Centre (UMMC) Medical Research Ethics Committees and the Ministry of Health Malaysia (No. 20191010-7917 and NMRR-19-3762-52429). Written informed consent was obtained from all subjects. The authors confirm that we have read the position of the Journal on issues regarding ethical publication and affirm that this work is consistent with those guidelines.

### Funding Sources

This project was funded by Global Parkinson’s Genetics Program (GP2). GP2 is funded by the Aligning Science Across Parkinson’s (ASAP) initiative and implemented by The Michael J. Fox Foundation for Parkinson’s Research (https://gp2.org). For a complete list of GP2 members, see https://gp2.org. Additionally, this project was supported by the Department of Medicine (DOM) Research Grant, Universiti Malaya (DOMRSF-2024-02/C5-1). Data used to prepare this article were obtained from the Accelerating Medicines Partnership® (AMP®) Parkinson’s Disease (AMP® PD) Knowledge Platform. For up-to-date information on the study, visit https://www.amp-pd.org. ACCELERATING MEDICINES PARTNERSHIP and AMP are registered service marks of the US Department of Health and Human Services. We thank the research participants for consenting to this study. The authors declare that there are no conflicts of interest relevant to this work.

### Financial Disclosures

EK has received honoraria from Elsevier for editorial duties. JHS has received honoraria from SK Chemical, Medtronics, Myungin Pharm, Handok, and Esai Korea, travel grants from the MDS and GP2, and research grants from the National Research Foundation of Korea, Seoul National University Hospital, and Seoul National University College of Medicine. HJK has received research grants from Seoul National University, Seoul National University Hospital, the Ministry of Health and Welfare, and the Ministry of Science and ICT, and holds stock ownership in PINE digital health Inc. BJ has received consultancies from Hanwha Pharm and SK Biopharm, and research grants from Zemvax & Kael. SK holds stock ownership in Genomic Therapeutics Pty Ltd and Prion OÜ, is an inventor for four inventions related to psoriasis and obesity drugs, has four patents related to psoriasis and obesity, is employed by Murdoch University, and has received five grants. KYM is employed by the The Hong Kong University of Science and Technology and received grant from Ng Teng Fong Charitable Foundation Limited, Lee Hysan Foundation, and DH Chen Foundation: NeuroCare Community Project (HK$30 million) (Date: 7/25 – 6/30) (PI: Ip N, Mok KY). IJKS has received research grants from the ASAP-GP2. MTP is employed by the University of Seville, has an honorary contract with QMUL, has received consultancies from UMEDEOR LTD, honoraria from the MJFF, and research grants from Parkinson’s UK. ZHF has a contract with the MJFF for Parkinson’s disease research and Data Tecnica International LLC. LML is employed by the Laboratories of Neurogenetics, National Institute on Aging, Bethesda, MD, USA, and the Institute of Neurogenetics, University of Luebeck, Luebeck, Germany, and has received faculty honoraria from the MDS. KRK is employed by NSW Health, serves on the Biogen Omaveloxolone Advisory Board, has received honoraria from the MDS, royalties from Oxford University Press, and research grants from the Medical Research Future Fund, Ainsworth 4 Foundation, and Lord Mayor Charitable Trust. SB is employed by Stellenbosch University, South Africa, and has received research grants from the South African Medical Research Council and the National Research Foundation of South Africa. JT is employed by the University of Luebeck, has received consultancies from Acurex Biosciences, serves on the Syngap Advisory Board, has received honoraria from the European Academy of Neurology, royalties from Elsevier, and research grants from the German Research Foundation Heisenberg, Else Kroener Fresenius Stiftung, and the MJFF. EMV has received honoraria for GP2-related activities and research grants from the Italian Ministry of Health, FRRB, and the Mariani Foundation. PH has received consultancies from EQT Life Sciences, Neuro.VC, and Singleton Biosciences, serves on the advisory boards of Alector Inc. and EURAC, holds stock in Alector Inc., Neuron23, and Sundance Biosciences, and holds several patents related to the diagnosis and treatment of neurodegenerative diseases. CK is employed by the University of Luebeck and University Hospital Schleswig-Holstein, Campus Luebeck, has received consultancies from Centogene and Biogen, speakers’ honoraria from Bial, royalties from Oxford University Press and Springer Nature, and research grants from DFG, MJFF, and ASAP. NEM is employed by Northwestern University, has received consultancies from Aspen Therapeutic, serves on the advisory board of the Parkinson’s Foundation, has received honoraria from the MDS, and research grants from NIH NINDS and ASAP-GP2. SYL is employed by the University of Malaya, Kuala Lumpur, Malaysia, has received consultancies from the Michael J. Fox Foundation (MJFF) for Parkinson’s research, and the Aligning Science Across Parkinson’s-Global Parkinson’s Genetics Program (ASAP-GP2), has received honoraria for participating as a Member of the Neurotorium Editorial Board, has received honoraria for lecturing/teaching from the International Parkinson and Movement Disorder Society (MDS) and Medtronic, has received stipends from the MDS as Chair of the Asian-Oceanian Section, and npj PD as Associate Editor, and has received research grants from MJFF. AAA has received a GP2 travel honorarium for the 2025 Investigator’s meeting and research grants from the GP2 PhD Fellowship (as a supervisor), the Ministry of Higher Education Malaysia, and the ALS Association USA. AHT is employed by the University of Malaya, Kuala Lumpur, Malaysia, serves as an associate editor for Elsevier, has received honoraria from the MDS, Eisai, Orion Pharma, and has received research grants from the MJFF and Global Parkinson’s Genetic Program (GP2). The other co-authors have no financial disclosures and conflict of interest to report.

