## Supplementary Material for "*GCH1* p.Ser80Asn Confers Risk for Parkinson’s Disease in East Asian Populations"

**This file includes:**

1. **Supplementary Methods …………………………………………….……………... 3**
2. **Supplementary Results ……………………………………….…………………….. 5**
3. **Supplementary Tables …….…………………………………………………….....7**

### Supplementary Methods

#### Genetic screening in proband and family

Peripheral blood (10mL) was collected from each subject. DNA was extracted using the FavorPrep™ Genomic DNA Extraction Maxi Kit (Favorgen, Taiwan) according to the manufacturer's protocol. The proband was screened via whole genome sequencing (WGS) via Global Parkinson's Genetics Program (GP2). Single nucleotide polymorphisms (SNPs), copy number variations (CNVs) and repeat expansions in known PD-related genes (Supplementary Table 2) were analysed. Candidate variants were validated via Sanger sequencing in the proband and his family members.

Additionally, the proband was screened via the SALSA® multiplex ligation-dependent probe amplification (MLPA) Probemix P051 kit (MRC Holland, Amsterdam, the Netherlands). (MLPA) assay for CNVs in known PD genes (*PRKN*, *PINK1*, *DJ-1*, *SNCA*, and *ATP13A2* [exons 2 and 9]). The data were analysed using the Coffalyzer software (https://www.mrcholland.com/technology/software/coffalyser-net).

#### Carrier frequency screening, association and haplotype analyses

In the GP2 WGS R11 datasets ($n=22{,}372_{PD}$; $n=3{,}578_{\text{other neurological disorders}}$; $n=8{,}826_{\text{Control}}$), populations across 11 ancestries were analyzed, including African American (AAC), African (AFR), Ashkenazi Jewish (AJ), Admixed American/Latin American (AMR), Complex Admixture History (CAH), Central Asian (CAS), East Asian (EAS), European (EUR), Middle Eastern (MDE), Finnish (FIN) and South Asian (SAS) (Supplementary Table 2).

The PLINK binary files of WGS data were processed using the standard quality control procedures of the GenoTools pipeline.[1] Samples with call rate >0.95, quality >20, read depth >5, and heterozygous allele balance between 0.25 and 0.75 were retained. Duplicated samples (kinship coefficient >0.354) and those with sex mismatches were removed. Ancestry prediction was performed via GenoTools[1] using the default reference panels from the 1000 Genomes Project, the Human Genome Diversity Project, and Ashkenazi Jewish datasets. Related individuals with second-degree or closer relatedness (kinship coefficient >0.0884) were excluded, leaving only the index cases for downstream analysis.

The genomic coordinates of 1kbp upstream and downstream *GCH1* p.Ser80Asn variant were obtained from the Ensembl database (hg38: chr14:54,901,425 - 54,903,425; https://www.ensembl.org). Variants within the region were extracted using PLINKv2.0, followed by annotation with ANNOVAR using reference datasets including refGene, dbNSFP v4.7a, ClinVar and gnomAD v4.1.1.

Additionally, GP2 WGS EAS cohort ($n=2{,}757_{PD}$ and $n=1{,}020_{\text{controls}}$) and independent EAS cohorts, including the EAS whole exome sequencing study (EAS-WES)[2] ($n=1{,}955_{PD}$ and $5{,}512_{\text{controls}}$), the Singaporean whole genome-based SG10K_Health project[3] ($n=9{,}770_{\text{controls}}$), and EAS controls from gnomAD (v4.1.0) ($n=22{,}431_{\text{controls}}$) were meta-analyzed to validate the findings. A two-tailed Fisher's exact test was used to compare allele frequencies of the *GCH1* p.Ser80Asn variant between cases and controls, and to estimate odds ratios (OR) with 95% confidence intervals (CI).

To determine whether the *GCH1* p.Ser80Asn variant carriers harbored a shared common haplotype, we phased chromosome 14 WGS data using Beagle v5.4[4] and searched for shared identical-by-descent (IBD) segments of length ≥2 cM among carriers using the hap-ibd function.[5] In addition, haplotypes surrounding the *GCH1* p.Ser80Asn variant were assessed using five flanking microsatellite markers (D14S991, D14S1057, D14S276, D14S52, and D14Skh3), previously described[6], spanning approximately 1.03 Mbp, with inter-marker distances ranging from 32 to 143 kbp. Genotyping was performed in nine Malaysian and Taiwanese carriers (Subjects 5-11, Subject 13 and his mother), Subject 13's unaffected father and brother (I:1 and II:1; Figure 1A), as well as five EAS controls. Moreover, SNP markers from GP2 WGS data of the carriers and 50 EAS controls were extracted across the same 1 Mbp region and used to supplement the microsatellite-based haplotype analysis and facilitate fine mapping. SNPs flanking the *GCH1* p.Ser80Asn variant were manually inspected, and the nearest upstream and downstream variants consistently observed in the non-reference state (heterozygous or homozygous alternate) among carriers were considered informative markers for defining the putative shared haplotype.

**Literature search for carriers of the *GCH1* p.Ser80Asn variant**

A PubMed search was conducted using the terms ("GCH1" OR "GTP cyclohydrolase 1") AND (dystonia OR "dopa-responsive dystonia" OR "DRD" OR "parkinson" OR "Parkinson's disease"), with no language restrictions for publications until 31st December 2025. All published cases carrying the *GCH1* p.Ser80Asn variant were extracted and summarized. Collaborators were contacted for clinical phenotyping data for identified *GCH1* p.Ser80Asn cases within the GP2 cohort.

### Supplementary Results

#### Clinical, neuroimaging and genetic findings of the proband and his family

#### Case description of the proband

The proband is a Chinese male (II:2, Figure 1A) from a non-consanguineous family who presented at age ranging 40-50 with progressive slowness in movements and gait impairment, without diurnal variation. On examination, he had bilateral moderate rigidity and bradykinesia, worse in the right upper and lower limbs. His gait was impaired with mild right foot dystonia characterised by right ankle inversion and toe extension, difficulty in initiation, short stride length and reduced arm swing. His eye movements and speech were normal. There were no tremor, other movement disorders, or pyramidal signs. His parkinsonism progressively worsened over several years, requiring increasing doses (LEDD: 950mg/day) of antiparkinsonian medications. By seven years post-diagnosis (at age ranging 50-60), he developed motor fluctuations ("wearing off" phenomenon lasting approximately three hours a day) and exhibited levodopa-induced dyskinesias.

At the most recent follow-up, the proband retained independence in activities of daily living but was unable to continue working as a driver. There was a progressive increase in disease severity, with his Clinical Impression of Severity Index for PD (CISI-PD) rising from 5 in 2019 to 12 in 2026. Between 2024 and 2026, his total MDS-UPDRS score increased slightly from 52 to 54, with a notable increase in non-motor burden (Part I: 3 to 8), and stable ON-state motor scores (Part III: 32 to 34). Current therapy consisted of levodopa–benserazide, piribedil, and selegiline (levodopa equivalent daily dosage [LEDD] 950 mg/day at 10 years of disease duration). Cognitive screening revealed mild impairment (Montreal Cognitive Assessment score 19/30; 9 years of formal education). Non-motor symptoms included REM sleep behaviour disorder (RBD), constipation, orthostatic hypotension, and pain. His medical history included essential hypertension and severe obstructive sleep apnoea (OSA) managed with continuous positive airway pressure (CPAP).

DaT-SPECT and 99mTc-TRODAT-1 SPECT/MRI fusion images (Figure 1B, v–vii) confirmed advanced nigrostriatal neurodegeneration with marked bilateral involvement of the putamen. This deficit was notably more pronounced on the left side, correlating with the patient's predominant right-sided motor findings. Axial SWI sequence images (Figure 1B, viii) revealed a loss of the left "swallow tail" sign and nigrosome-1 hyperintensity.

#### Case description of the proband's mother

His mother (I:2, Figure 1A) was incidentally found to have mild parkinsonism at age ranging 70-80 in 2024, characterized by mild bradykinesia in all limbs and slight rigidity on reinforcement, more notable on the right side. Gait was slow but with good stride length and arm swing. She did not have tremor, dystonia, other movement disorders, or pyramidal signs. Longitudinal assessment from 2024 to 2026 revealed progressive decline, with her total MDS-UPDRS score increasing from 32 to 49, primarily driven by worsening motor examination findings (Part 3: 23 to 34). By 2026, her CISI-PD was 5, and cognitive screening showed mild impairment (MoCA 21/30; six years of formal education). Over the two-year follow-up, she continued to be fully independent and employed in manual work. She was treated with low-dose levodopa–benserazide (LEDD 100 mg/day at 2 years of disease duration).

Motor and non-motor features of both proband and his mildly affected mother are summarised in Table 1 with accompanying videos in the Supplementary Materials. DaT-SPECT and 99mTc-TRODAT-1 SPECT/MRI fusion images (Figure 1B, i–iii) demonstrated nigrostriatal neurodegeneration characterized by mild, primarily unilateral loss. In contrast to the functional imaging, structural assessment via axial SWI sequence (Figure 1B, iv) showed a preserved bilateral "swallow tail" sign (white arrows).

**Haplotype analyses**

As the variant was exclusively observed in individuals of EAS ancestry within the GP2 WGS cohort, haplotype analyses were performed to evaluate whether carriers shared a common ancestral haplotype. However, no shared disease-associated haplotype was detected among the 10 *GCH1* p.Ser80Asn carriers following Beagle phasing. Consistent with this result, neither microsatellite marker analysis nor inspection of flanking SNP genotype patterns across the same ~1 Mbp region identified a common disease-associated haplotype among the *GCH1* p.Ser80Asn carriers (Supplementary Table 3). A schematic diagram to illustrate the segregation of microsatellite and SNP marker alleles in the Malaysian Chinese family was shown in Supplementary Figure 1.

**Supplementary Figure 1: Haplotype analyses surrounding the *GCH1* p.Ser80Asn variant in a Malaysian Chinese family using microsatellite and SNP markers.**

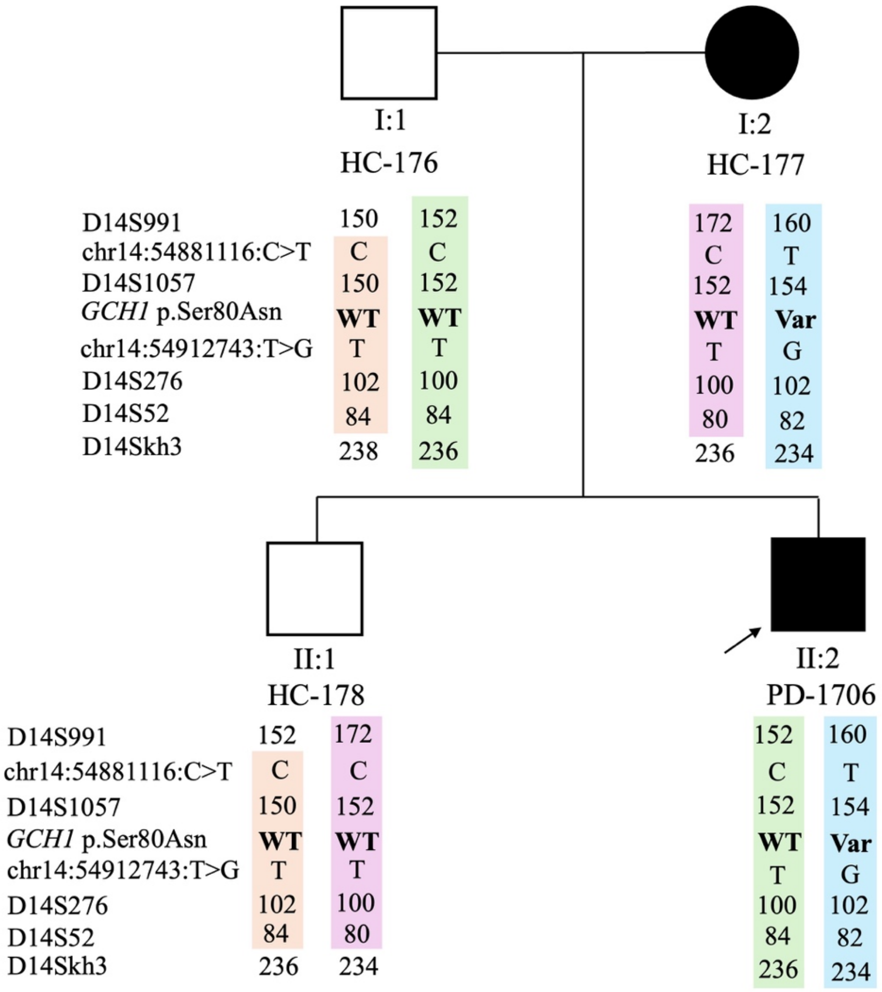


Schematic representation of the segregation of microsatellite and SNP marker alleles flanking the *GCH1* p.Ser80Asn variant within the Malaysian Chinese family. Var=Variant; WT=Wildtype.

**Supplementary Tables**

**Supplementary Table 1: Demographics of the multi-ancestral GP2 R11 cohorts with whole genome sequencing data.**

| Genetic ancestry | Number of subjects | | | *GCH1* p.Ser80Asn variant carrier, *n* |
|---|---|---|---|---|
| | PD, *n* [1] | Patients with other neurological disorders, *n* [2] | Control, *n* [3] | |
| AAC | 245 | 6 | 157 | 0 |
| AFR | 1,495 | 33 | 2,063 | 0 |
| AJ | 1,190 | 52 | 234 | 0 |
| AMR | 907 | 8 | 300 | 0 |
| CAH | 234 | 25 | 117 | 0 |
| CAS | 684 | 125 | 954 | 0 |
| EAS | 2,757 | 474 | 1,020 | 10<br>(9 PD and 1 control) |
| EUR | 13,416 | 2,761 | 2,678 | 0 |
| FIN | 20 | 5 | 4 | 0 |
| MDE | 945 | 26 | 967 | 0 |
| SAS | 479 | 63 | 332 | 0 |
| **Total** | **22,372** | **3,578** | **8,826** | **10** |

AAC=African American; AFR=African; AJ=Ashkenazi Jewish; AMR=Admixed American/Latin American; CAH=Complex Admixture History; CAS=Central Asian; EAS=East Asian; EUR=European; FIN=Finnish; MDE=Middle Eastern; *n*=Number; SAS=South Asian.

[1] The "PD" category includes individuals with PD from i) unselected PD case/control cohorts, ii) monogenic recruitment, i.e., targeted recruitment of individuals in which a monogenic cause of disease is more likely based on a young age at onset <50 years and/or a positive family history of

PD), and iii) genetically enriched cohorts, i.e., targeted recruitment of carriers of pathogenic variants in *GBA1*, *LRRK2*, or *SNCA*.

[2] The “Other neurological disorders” category summarizes individuals with phenotypes other than classical PD, including atypical forms of parkinsonism (e.g., multisystem atrophy, progressive supranuclear palsy, corticobasal syndrome, and dementia with lewy bodies) and other neurodegenerative phenotypes (e.g., Alzheimer’s disease, mild cognitive impairment, other forms of dementia).

[3] The “control” category summarizes all individuals without PD or other phenotypes listed above, including healthy controls, population cohorts, and healthy family members of individuals with PD or parkinsonism.

**Supplementary Table 2: PD and neurological disorder-related genes screened in the proband of the Malaysian Chinese family via whole genome sequencing.**

| **A. PD and neurological disorder-related genes screened for single nucleotide polymorphism and copy number variations** | | | | |
|---|---|---|---|---|
| *AFG3L2* | *EPM2A* | *NR4A2* | *QDPR* | *SYNJ1* |
| *ATP13A2* | *FA2H* | *OPA1* | *RAB39B* | *TAF1* |
| *ATP1A3* | *FBXO7* | *OPA3* | *SCP2* | *TBK1* |
| *ATP6AP2* | *FTL* | *PANK2* | *SLC20A2* | *TH* |
| *ATP7B* | *GBA* | *PARK7* | *SLC25A4* | *TUBB4A* |
| *C19orf12* | *GCH1* | *PDE8B* | *SLC25A46* | *TWNK* |
| *CHCHD10* | *GLB1* | *PDGFB* | *SLC30A10* | *VAC14* |
| *CHCHD2* | *GRN* | *PDGFRB* | *SLC39A14* | *VCP* |
| *COASY* | *JAM2* | *PINK1* | *SLC6A3* | *VPS13A* |
| *CP* | *LRRK2* | *PLA2G6* | *SMPD1* | *VPS13C* |
| *CSF1R* | *LYST* | *POLG* | *SNCA* | *VPS35* |
| *DCAF17* | *MAPT* | *PRKN* | *SPG11* | *WDR45* |
| *DCTN1* | *MYORG* | *PRKRA* | *SPG7* | *XPR1* |
| *DNAJC12* | *NPC1* | *PTRHD1* | *SPR* | *ZFYVE26* |
| *DNAJC6* | *NPC2* | *PTS* | *SQSTM1* | |
| **B. PD and neurological disorder-related genes screened for repeat expansions** | | | | |
| *AFF2* | *ATXN7* | *DMPK* | *NIPA1* | *RFC1-AAGGG* |
| *AR* | *AYXN8OS* | *FGF14* | *NOP56* | *RFC1-ACAGG* |
| *ARX_1* | *BEAN1* | *FMR1* | *NOTCH2NLC* | *SAMD12* |
| *ARX_2* | *C9ORF72* | *FXN* | *NUTM2B-AS1* | *SOX3* |
| *ATN1* | *CACNA1A* | *GIPC1* | *PABPN1* | *STARD7* |
| *ATXN1* | *CBL* | *GLS* | *PHOX2B* | *TBP* |
| *ATXN10* | *CNBP* | *HTT* | *PPP2R2B* | *TNRC6A* |
| *ATXN2* | *DAB1* | *JPH3* | *PRDM12* | *YEATS2* |
| *ATXN3* | *DIP2B* | *MARCHF6* | *RAPGEF2* | *ZIC2* |

**Supplementary Table 3: Haplotypes analyses surrounding the *GCH1* p.Ser80Asn variant in East Asian carriers and controls using microsatellite and SNP markers.**

| **Subject** | | **5** | **6** | **7** | **8** | **9** | **10** | **11** | **12** | **13 - Proband** | **13's mother** | **C1** | **13's father** | **13's brother** | **C2** | **C3** | **C4** | **C5** | **C6** | **C7** | **C8** |
|---|---|---|---|---|---|---|---|---|---|---|---|---|---|---|---|---|---|---|---|---|---|
| **Microsatellite or SNP markers** | **Chr position (hg38)** | | | | | | | | | | | | | | | | | | | | |
| D14S991 | 54757307 | 144/150 | 144/152 | 144/152 | 144/150 | 144/152 | 132/132 | 152/150 | NA | 152/160 | 172/160 | NA | 150/152 | 152/172 | 150/160 | 134/150 | 156/160 | 152/160 | 134/134 | NA | NA |
| chr14:54881116: C>T | 54881116 | C/T | C/T | T/T | C/T | C/T | C/T | C/T | C/T | C/T | C/T | C/T | C/C | C/C | C/C | C/C | C/C | C/C | C/C | C/T | T/T |
| D14S1057 | 54900756 | 152/154 | 152/154 | 152/154 | 152/154 | 166/154 | 162/154 | 152/154 | NA | 152/154 | 152/154 | NA | 150/152 | 150/152 | 158/162 | 152/150 | 152/156 | 148/152 | 148/164 | NA | NA |
| *GCH1* p.Ser80Asn | 54902425 | WT/Var | | | | | | | | | | | WT/WT | | | | | | | | |
| chr14:54912743: T>G | 54912743 | T/G | T/G | T/G | T/G | T/G | T/G | T/G | T/G | T/G | T/G | T/G | T/T | T/T | T/T | T/T | T/T | T/T | T/T | T/G | T/G |
| D14S276 | 55216298 | 94/102 | 94/102 | 94/106 | 100/102 | 94/94 | 94/94 | 94/102 | NA | 100/102 | 100/102 | NA | 102/100 | 102/100 | 104/94 | 102/106 | 102/102 | 100/102 | 100/106 | NA | NA |
| D14S52 | 55758620 | 84/86 | 84/86 | 82/86 | 74/86 | 82/86 | 80/82 | 72/82 | NA | 84/82 | 80/82 | NA | 84/84 | 84/80 | 74/74 | 82/84 | 80/80 | 86/88 | 86/88 | NA | NA |
| D14Skh3 | 55790598 | 236/234 | 226/238 | 230/238 | 230/238 | 228/230 | 230/236 | 228/230 | NA | 236/234 | 236/234 | NA | 238/236 | 236/234 | 228/230 | 230/236 | 234/242 | 228/230 | 236/234 | NA | NA |

Genotype patterns of the flanking microsatellite and informative SNP markers surrounding the GCH1 p.Ser80Asn variant in carriers and East Asian (EAS) controls are shown. Two flanking SNPs, chr14:54881116:C>T (upstream) and chr14:54912743:T>G (downstream), consistently observed in the non-reference state (heterozygous or homozygous alternate) among carriers were selected as informative markers. C=Control; SNP=Single nucleotide polymorphism; Var=Variant; WT=Wildtype.
